# Differentials and determinants of immunization coverage among children aged 12-36 months in India: Analysis of nationally-representative, population-based survey data

**DOI:** 10.1101/2021.08.26.21262652

**Authors:** Eram Naaz, Saddaf Naaz Akhtar, Manzoor Ahmad Malik, Jalandhar Pradhan

**Author notes:** **Corresponding author:** Eram Naaz, Student at Department of Humanities and Social Sciences, National Institute of Technology (NIT), Rourkela, Odisha.

## Abstract

**Introduction:** Immunization coverage among children is still a major public health concern in India and other low-middle income countries. Low coverage likely risks the health of children and therefore impacts their overall growth. We therefore examined the immunization coverage rates among children aged 12-36 months in India and its states. We also explored the associated factors affecting immunization coverage among children aged 12-36 in India.

**Methods:** We used data from 75^th^ round of the National Sample Survey Organizations (NSSO), collected from July 2017 to June 2018. The analytical sample of children aged 12-36 months information cases was 15887. Immunization coverage rates of India and its states were calculated. We evaluated the immunization coverage rate by background characteristics in India and its states. We performed multinomial logistic regression analysis to estimate the factors associated with the immunization coverage in India.

**Results:** About 61.95% were fully immunized, 35.62% were partially immunized and the rest 2.43% had not received any vaccine. The children aged 21-28 months (0.50; p<0.01) & 29-36 months (0.35; p<0.01), belonging to North-Eastern regions (0.60; p<0.01) were found to be significantly less likely to receive fully immunization. Likewise, urban children are (1.26; p<0.1) found to be more likely to receive fully immunization. The lowest fully immunization has been seen in Daman and Diu (1.73%) followed by Nagaland (6.93%), NCT Delhi (34.71%), and Tripura (46.33%).

**Conclusions:** Child immunization is a key public health concern and vital challenge to be addressed. Socio-economic characteristics play a key role in immunization coverage. Therefore necessary policy measures must be taken to address the challenges of poor immunization coverage and its impact on health and wellbeing of children

## Introduction

Immunization is an essential part of primary health care and a basic human right. Immunization is one of the most beneficial public health measures of the twentieth century for humans [1]. Immunization means protection, an infant from contracting a variety of diseases. Many of these diseases are easily passed from child to child and can cause severe health problems, they have the capability of being fatal [2]. Immunization can prevent him/her from major disease or bundle of the diseases through vaccine, some of the disease such as polio, hepatitis B, measles, tetanus, pertussis, diphtheria etc [3]. Immunizations have prevented epidemics of once-common infectious diseases like measles, mumps, and whooping cough over the past year [4].

Prior to 2010, child mortality has been declined by one fourth at a global level [5]. Globally, the immunization coverage has been dropped from 86% in 2019 to 83% in 2020 [6]. While more children are being immunized than ever before which is a great stride that have been made in immunization in worldwide [5]. By protecting children against unsafe diseases, vaccines can also play a vital role in ending preventable child deaths [7]. Childhood immunization is an effective tool for preventing illness and ensuring a child’s survival, as well as providing opportunities for children to succeed [8]. Expanding childhood immunization efforts has helped many countries achieve significant achievements in recent decades, such as the eradication of vaccine-preventable diseases and improved population herd immunity [9]. Unfortunately, many children remain unvaccinated or under vaccinated, and vaccine-preventable diseases are still a leading cause of morbidity and mortality, especially in low- and middle-income countries [10]. Although, global campaigns aimed at ensuring that all children receive vaccines have increased in number and strength, attracting broad support from the international public health community. On the ground, however, progress has slowed in recent years, especially among the most vulnerable and difficult-to-reach populations [11].

Childhood immunisation campaigns are being refocused in order to prevent health risks and deaths [9]. Childhood immunisation can help achieve Sustainable Development Goals (SDG) 3: to ensure safe lives and promote wellness for all people of all ages, especially given emerging global health priorities and needs [12]. Childhood immunisation is a central component of SDG target 3.2, which focuses on avoiding preventable deaths in new-borns and children under the age of five, and SDG target 3.8, which seeks to achieve equitable access to vital medicines and vaccines. Full immunisation coverage, for example, is one of the proposed metrics for the health-related SDGs as a way to measure progress toward universal health coverage [12].

Moreover, social disparities in vaccination uptake can thwart global efforts to reduce disease burden in low- and middle-income countries, since children from low- and middle-income families are more likely to contract infectious diseases [9]. There is a wide body of literature dedicated to factors that affect vaccine adoption because of the millions of preventable infant deaths that occur each year in low- and middle-income countries as a result of inadequate or total lack of vaccination [13]. Vaccination coverage is influenced by a variety of variables, including income distribution, maternal education, residency, infant gender, and poverty. Children’s vaccination coverage is also uneven. According to studies of the health status of precarious households, migrant and refugee children, and Roma children, a large number of infants are not receiving the vaccines that they need [14]. It is important to invest in health-care professionals that can administer vaccines and provide updates to families [10].

In India, socio-economic disparities in health care are the major issues [15]. Vaccination disparities among children are still a major public health concern around the world [13]. Children’s malnutrition is a serious public health problem that has a negative impact on infant and young child mortality, morbidity, and life expectancy [16]. The Indian government’s Universal Immunization Program (UIP) and Reproductive and Child Health Program (R&CHP) have improved childhood vaccination rates and minimised socioeconomic inequalities [17]. The Indian government funds all vaccine doses recommended in the UIP and provides them free of charge to all children in public healthcare facilities [18]. In low-income states, where more than half of India’s children under the age of five live, the private health sector’s contribution to childhood vaccination is limited, and the government administers the majority of vaccinations [19].

However, there is a need of the study basically, to raise and sustain immunization coverage levels in India. According to WHO reports, childhood vaccines prevent 2–3 million untimely deaths in children per year, with an additional 1.5 million deaths likely to be avoided with increased vaccination coverage [9]. So, it is important to see whether there any differential exists in immunization coverage rates among children aged 12-36 in India and how it varies from states to states? And what are the determinants that are associated with the immunization coverage among children aged 12-36 in India?

Therefore, our study aims to examine the immunization coverage rates among children aged 12-36 months in India and its states. And then determine the associated factors affecting immunization coverage among children aged 12-36 in India.

## Data source

The present study has used the data from 25^th^ schedule of the 75^th^ round of the National Sample Survey Organizations (NSSO), collected from July 2017 to June 2018. The NSSO is a public organization since 1950 under the Ministry of Statistics and Programme Implementation (MOSPI) of the Government of India.

## Analytical approach

The analytical sample of children aged 12-36 months information cases was 15887.

## Methodology

These variables of immunization coverage status were dichotomous variable. In the questionnaire, it has been asked to the respondent that whether the children had ever received immunization during the last 365 days at the time of survey. *Immunization (yes) is taken as “1” and No-immunization (no) as “0”*.

### Dependent variable

Thus, the dependent variable here is defined as Immunization coverage which is consists of three categories are:

i. ***No Immunization coverage:*** Those children who has not received any vaccination or immunization services in the last 365 days at the time of the survey.
ii. ***Partly immunization coverage:*** Those children who has received at least one or up to seven doses vaccination or immunization services in the last 365 days at the time of the survey.
iii. ***Fully immunization coverage:*** Those children who has taken all eight vaccination or immunization services in the last 365 days at the time of the survey.

### Independent variables

The control variables focus on socio-demographic, economic characteristics among children aged 12-36 months in India. The socio-demographic and economic characteristics include the children’s age groups, place of residence, regions, wealth quintiles, religion, social-groups (caste groups) in the last 365 days prior to the survey.

### Analysis

The primary analysis used univariate, bivariate cross-tabulations and percentages were calculated by socio-demographic, economic characteristics among children aged 12-36 months in India. The present study has evaluated the immunization coverage rate by background characteristics in India and its states. And then, the study has used multinomial logistic regression analysis to estimate the factors associated with the immunization.

## Results

### Characteristics of analytical sample

Table 1 shows the sample profiles by socio-demographic and economic background characteristics among the children in India. The percentage of children age 21-28 months is highest (45.27%) compared to other age groups. There are (51.62%) male children aged 12-36 months and (48.38%) female children aged 12-36 months. The percentage of rural children is found to be (62.01%) which is higher than urban children (37.99%). The percentage of regions in central is found to be (21.67%) which is higher than the other region groups, and the lower in north-eastern region found to be (11.61%) among children aged 12-36 months. Hindu’s children are showing highest (71.73%) compared to other religion groups. Non-Hindu’s children are showing lower (28.27%) in comparing to the Hindu’s children and the others religion groups. The percentage of caste in which other backward caste is showing highest (41.76%) than the other caste groups, and Schedule Caste/Schedule Tribes is found to be (33.1%) which is lower than the other backward caste groups.

**Table 1.**
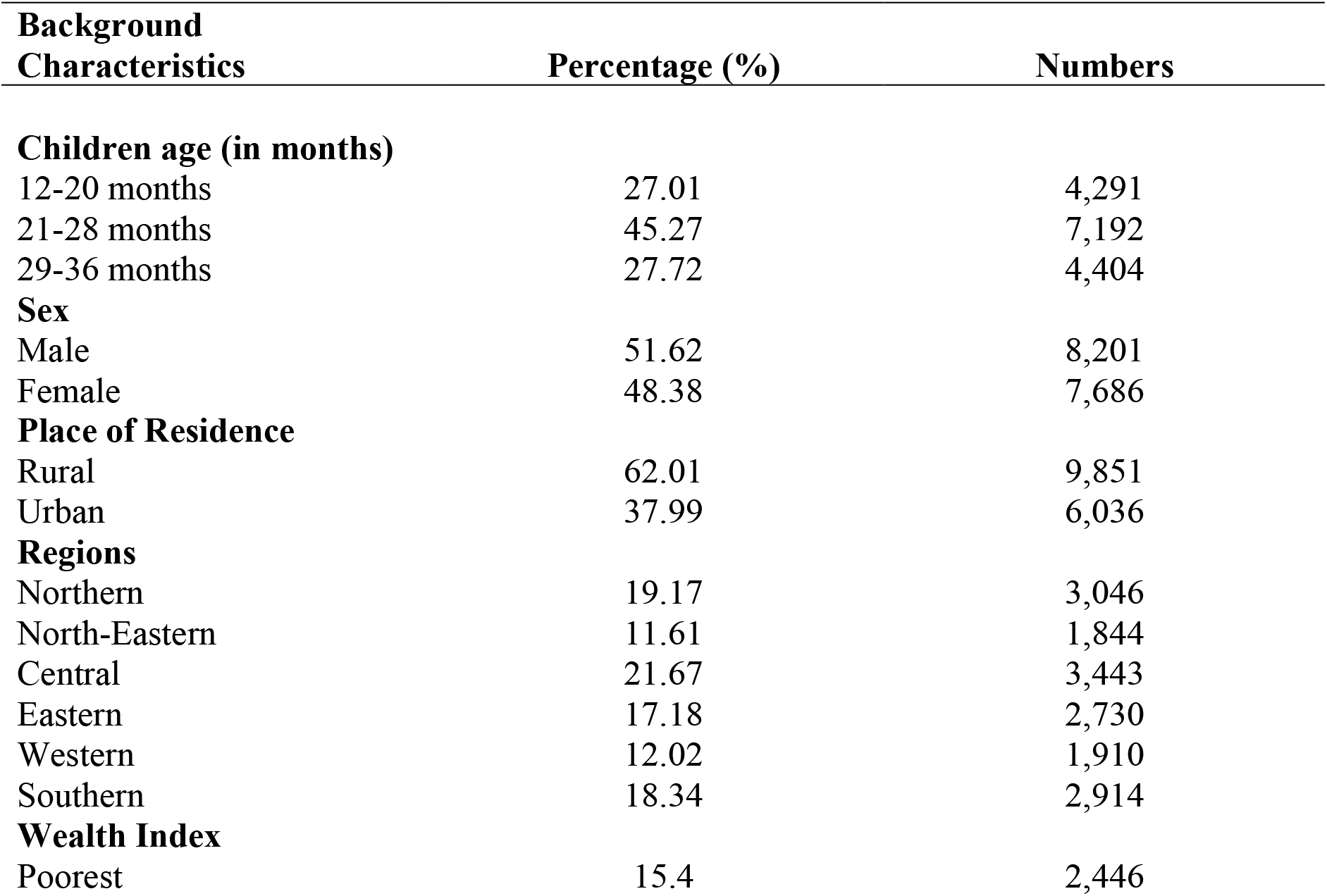
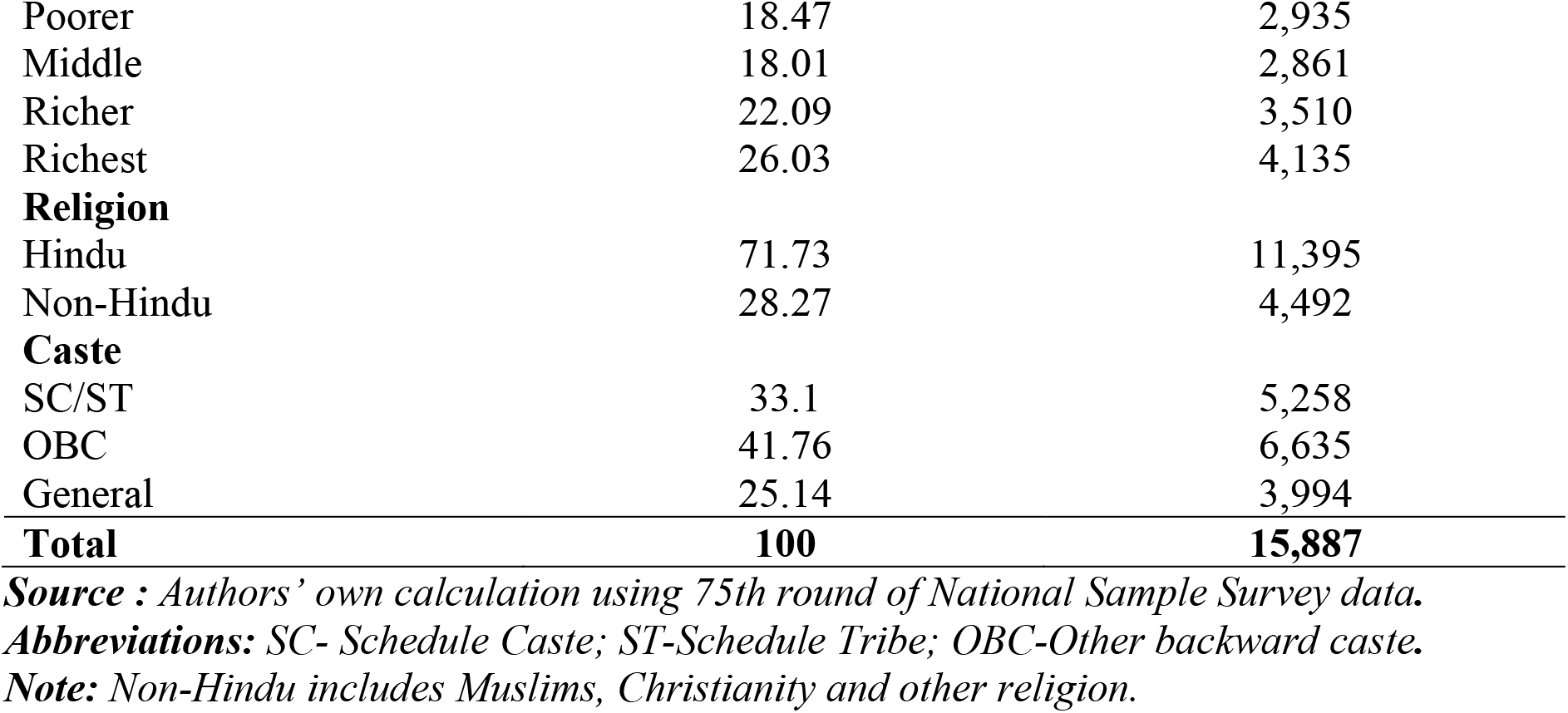
Sample profile by socio-demographic and economic background characteristics among under-five children in India (n=15,887).

| Background Characteristics | Percentage (%) | Numbers |
| --- | --- | --- |
| **Children age (in months)** |  |  |
| 12-20 months | 27.01 | 4,291 |
| 21-28 months | 45.27 | 7,192 |
| 29-36 months | 27.72 | 4,404 |
| **Sex** |  |  |
| Male | 51.62 | 8,201 |
| Female | 48.38 | 7,686 |
| **Place of Residence** |  |  |
| Rural | 62.01 | 9,851 |
| Urban | 37.99 | 6,036 |
| **Regions** |  |  |
| Northern | 19.17 | 3,046 |
| North-Eastern | 11.61 | 1,844 |
| Central | 21.67 | 3,443 |
| Eastern | 17.18 | 2,730 |
| Western | 12.02 | 1,910 |
| Southern | 18.34 | 2,914 |
| **Wealth Index** |  |  |
| Poorest | 15.4 | 2,446 |
| Poorer | 18.47 | 2,935 |
| Middle | 18.01 | 2,861 |
| Richer | 22.09 | 3,510 |
| Richest | 26.03 | 4,135 |
| **Religion** |  |  |
| Hindu | 71.73 | 11,395 |
| Non-Hindu | 28.27 | 4,492 |
| **Caste** |  |  |
| SC/ST | 33.1 | 5,258 |
| OBC | 41.76 | 6,635 |
| General | 25.14 | 3,994 |
| **Total** | **100** | **15,887** |

**Source :** Authors' own calculation using 75th round of National Sample Survey data.
**Abbreviations:** SC- Schedule Caste; ST-Schedule Tribe; OBC-Other backward caste.
**Note:** Non-Hindu includes Muslims, Christianity and other religion.

#### Immunization coverage patterns in India

Table 2 shows the immunization coverage pattern by socio-demographic and economic background characteristics among the children aged 12-36 months in India. Of the total included children (N=15,887), (61.95%) were fully immunized, (35.62%) were partially immunized and the rest (2.43%) had not received any vaccine. Around (36.31 %) male children were partially immunized than female children (34.9%) whereas, (62.93%) female children are fully immunized compared to the male children (61.01%). Urban children have received (3.16%) lesser partial immunization services than rural children, on the other hand urban children have (4.71%) higher fully immunization services than rural children.

**Table 2.** Immunization coverage pattern by socio-demographic and economic background characteristics among the children aged 12-36 months in India (n=15,887).

| **Background Characteristics** | **No immunization (%)** | **Partial immunization (%)** | **Full Immunization (%)** |
| --- | --- | --- | --- |
| **Children age (in months)** |  |  |  |
| 12-20 months | 0.71 | 45.34 | 53.95 |
| 21-28 months | 2.67 | 34.98 | 62.35 |
| 29-36 months | 4.04 | 25.2 | 70.76 |
| **Sex** |  |  |  |
| Male | 2.68 | 36.31 | 61.01 |
| Female | 2.17 | 34.9 | 62.93 |
| **Place of Residence** |  |  |  |
| Rural | 2.73 | 36.47 | 60.81 |
| Urban | 1.62 | 33.31 | 65.07 |
| **Regions** |  |  |  |
| Northern | 3.26 | 34.22 | 62.52 |
| North-Eastern | 3.8 | 43.2 | 53 |
| Central | 1.81 | 38.58 | 59.6 |
| Eastern | 2.61 | 38.47 | 58.92 |
| Western | 2.22 | 36.99 | 60.8 |
| Southern | 2.32 | 25.93 | 71.74 |
| **Wealth Index** |  |  |  |
| Poorest | 2.75 | 40.07 | 57.18 |
| Poorer | 3.15 | 38.17 | 58.68 |
| Middle | 2.03 | 34.65 | 63.32 |
| Richer | 2.36 | 33.66 | 63.97 |
| Richest | 1.81 | 31.45 | 66.74 |
| **Religion** |  |  |  |
| Hindu | 2.52 | 36.28 | 61.2 |
| Non-Hindu | 2.12 | 33.3 | 64.59 |
| **Caste** |  |  |  |
| SC/ST | 2.31 | 37.41 | 60.28 |
| OBC | 2.44 | 35.53 | 62.03 |
| General | 2.59 | 33.19 | 64.22 |
| **Total** | **2.43** | **35.62** | **61.95** |

**Source :** Authors' own calculation using 75th round of National Sample Survey data.
**Abbreviations:** SC- Schedule Caste; ST-Schedule Tribe; OBC-Other backward caste.
**Note:** Non-Hindu includes Muslims, Christianity and other religion.

#### Immunization coverage pattern among states of India

The Figure 1 shows the no immunization coverage pattern in 36 Indian states. The highest no immunization coverage has been found in Chandigarh (18.54%) followed by Arunachal Pradesh (8.26%), Tripura (6.63%), and Manipur (5.98%). Whereas Andaman & Nicobar Island, Dadar & Nagar Haveli, Daman & Diu, Goa, Kerala, Lakshadweep, Meghalaya, Puducherry, Sikkim and Uttarakhand showed zero no-immunization coverage. However, the Figure 2 shows the partially immunization coverage pattern in 36 states of India, where the children aged 12-36 months belonging to Daman & Diu (98.27%) has the highest partially immunization followed by Nagaland (89.2%), NCT of Delhi (65.27%) and Puducherry (51.29%). On the other hand, the lowest partially immunization coverage has been depicted in Manipur (8.9%) followed by Mizoram (15.45%), Telangana (18.81%), and Himachal Pradesh (18.89%). Despite of that, the Figure 3 shows the fully immunization coverage pattern among 36 states of India. The highest fully immunization coverage has been found in Manipur (85.12%) followed by Mizoram (83%), Telangana (80.93%), Haryana (79.87%), and Kerala (78.73%). But the lowest fully immunization has been seen in Daman and Diu (1.73%) followed by Nagaland (6.93%), NCT Delhi (34.71%), and Tripura (46.33%).

**Figure 1.**
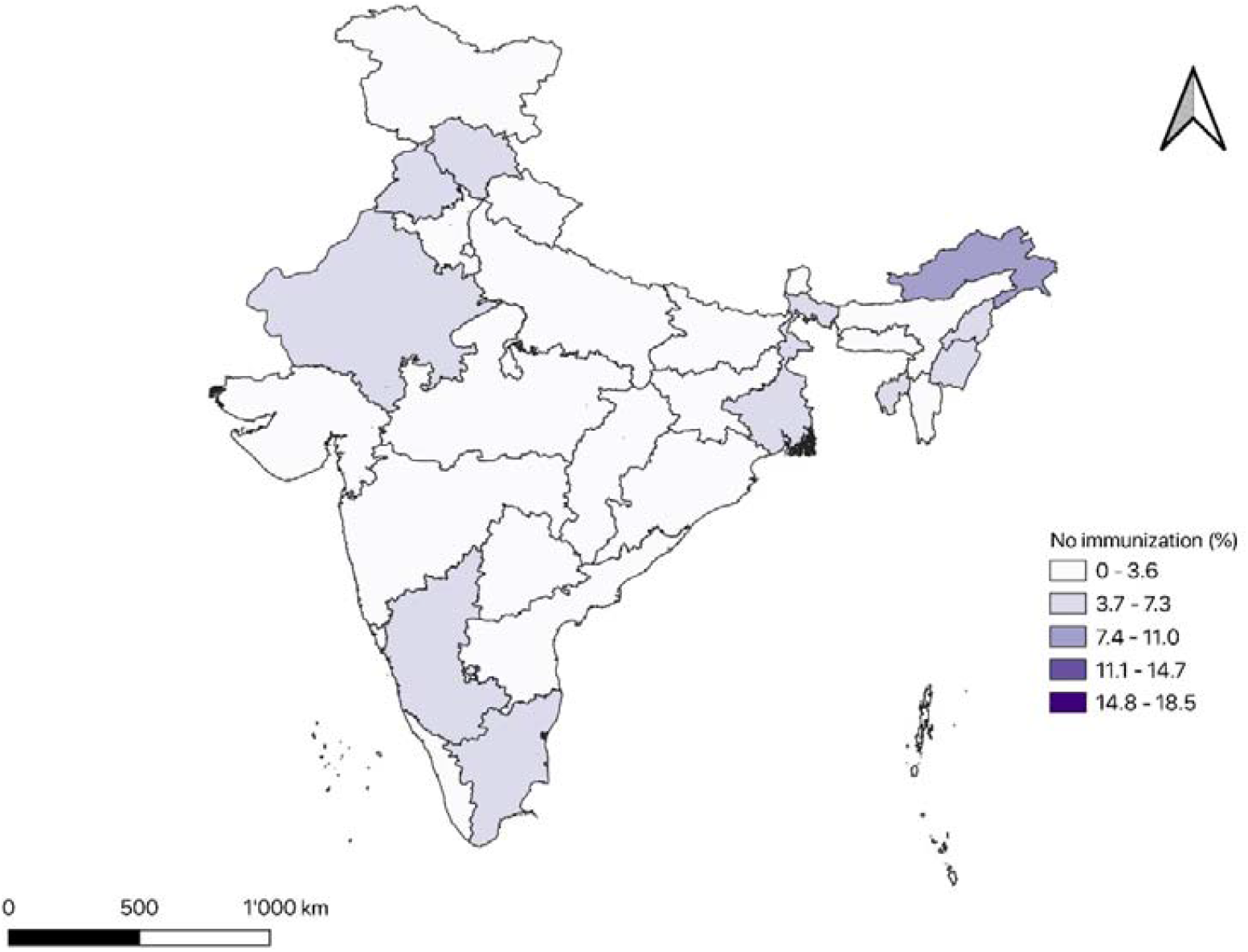
No-immunization coverage pattern among the children aged 12-36 months in India. **Note:** The map is showing the no-immunization coverage pattern among the children aged 12-36 months in 36 states of India (%).

**Figure 1.**
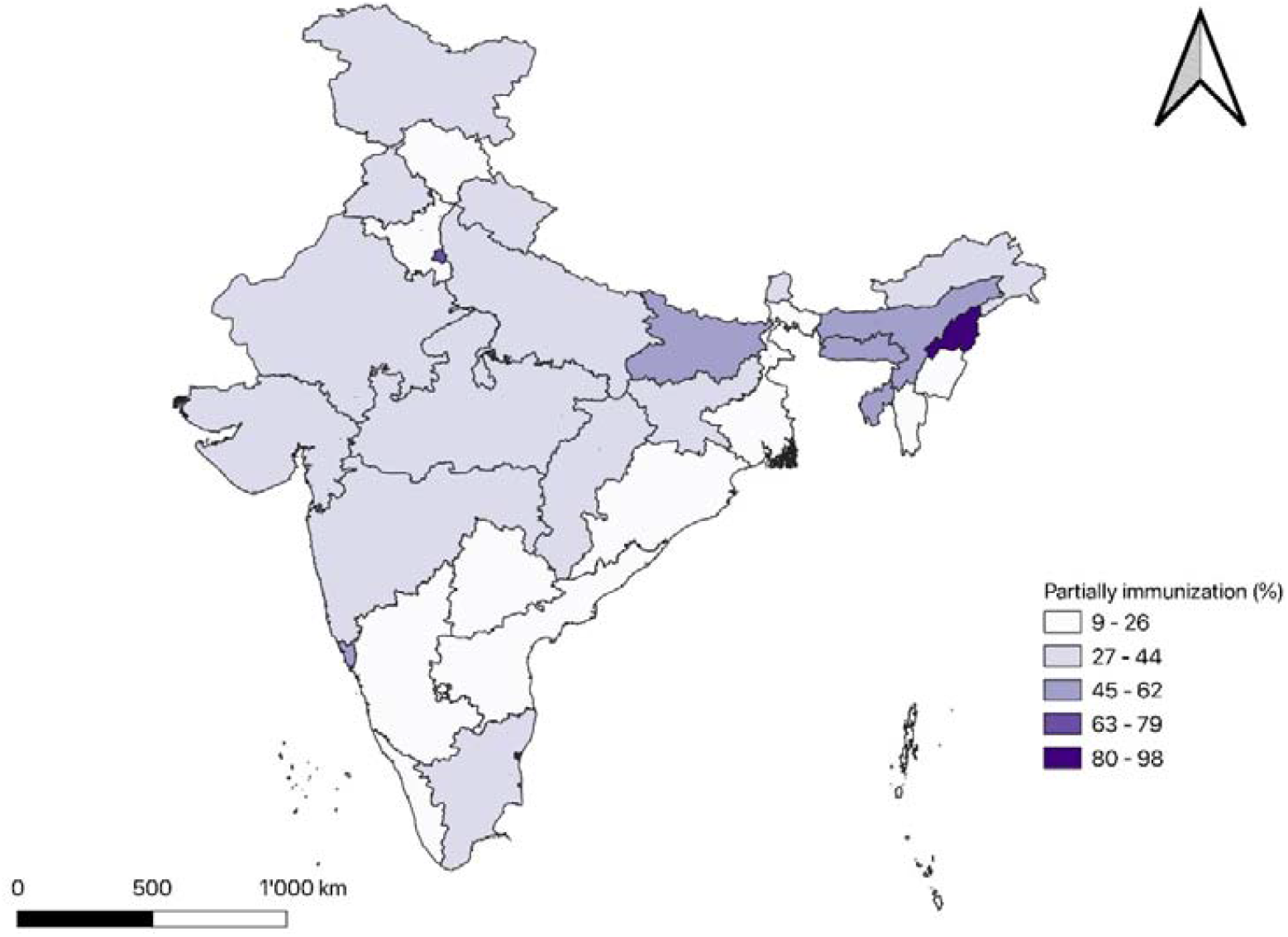
Partially immunization coverage pattern among the children aged 12-36 months in India. **Note:** The map is showing the partial immunization coverage pattern among the children aged 12-36 months in 36 states of India (%).

**Figure 2.**
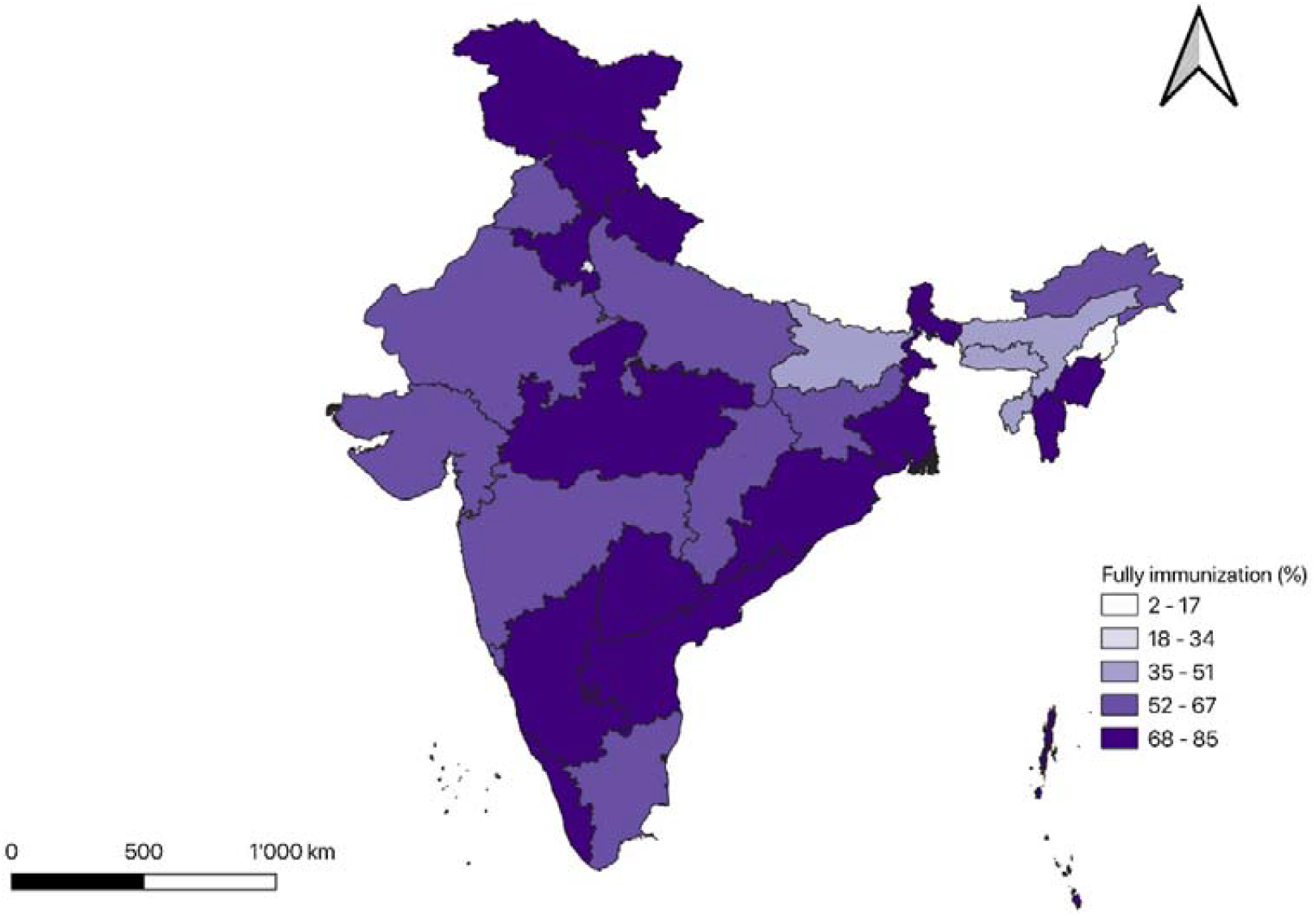
Fully immunization coverage pattern among the children aged 12-36 months in India. **Note:** The map is showing the fully immunization coverage pattern among the children aged 12-36 months in 36 states of India (%).

#### Factors affecting immunization coverage in India

Table 3 presents the results of multinomial logit regression, examining the associated factors affecting immunization coverage among the children in India, after adjusting the role of demographic, socioeconomic background characteristics. The foremost finding of this analysis is that the children aged 21-28 months are found to be significant and less likely to receive partially immunization compared to children aged (0.29; p<0.01) 12-20 months & (0.14; p<0.01) 29-36 months compared to 12-20 months. Children living in urban areas (1.24; p<0.1) are more likely to receive partially immunization than in rural areas. Poorer children (0.64; p<0.01) are significantly less likely to receive partial immunization compared to poorest children.

**Table 3.**
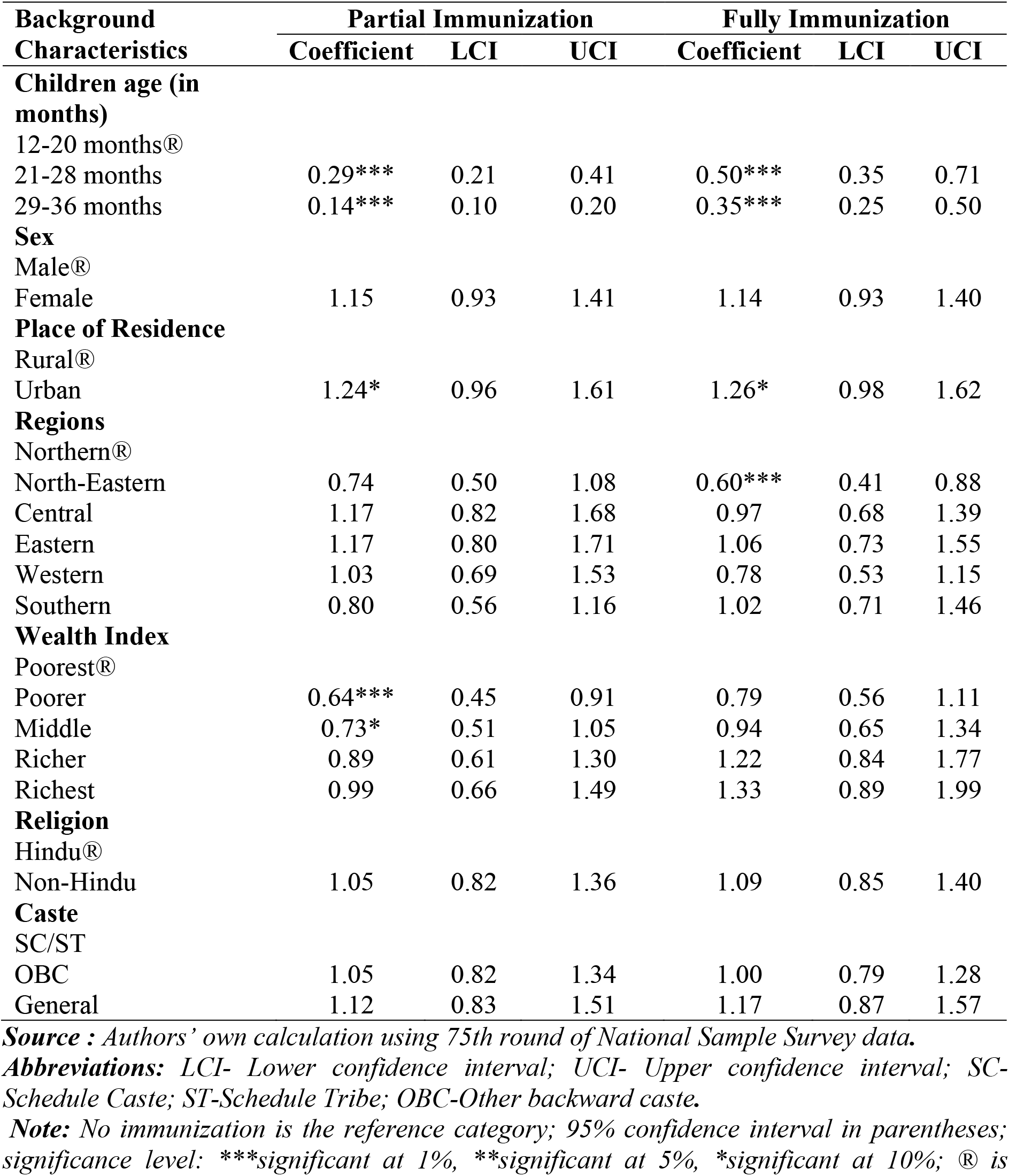
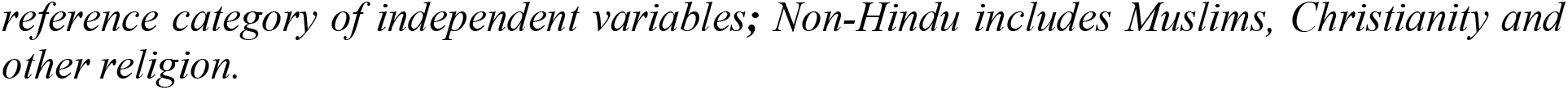
Results of Multinomial Logit Regression: Predictors of Immunization coverage among children in India, 2017-18 (n=15,887).

| **Background Characteristics** | **Partial Immunization** |  |  | **Fully Immunization** |  |  |
| --- | --- | --- | --- | --- | --- | --- |
|  | **Coefficient** | **LCI** | **UCI** | **Coefficient** | **LCI** | **UCI** |
| **Children age (in months)** |  |  |  |  |  |  |
| 12-20 months® |  |  |  |  |  |  |
| 21-28 months | 0.29*** | 0.21 | 0.41 | 0.50*** | 0.35 | 0.71 |
| 29-36 months | 0.14*** | 0.10 | 0.20 | 0.35*** | 0.25 | 0.50 |
| **Sex** |  |  |  |  |  |  |
| Male® |  |  |  |  |  |  |
| Female | 1.15 | 0.93 | 1.41 | 1.14 | 0.93 | 1.40 |
| **Place of Residence** |  |  |  |  |  |  |
| Rural® |  |  |  |  |  |  |
| Urban | 1.24* | 0.96 | 1.61 | 1.26* | 0.98 | 1.62 |
| **Regions** |  |  |  |  |  |  |
| Northern® |  |  |  |  |  |  |
| North-Eastern | 0.74 | 0.50 | 1.08 | 0.60*** | 0.41 | 0.88 |
| Central | 1.17 | 0.82 | 1.68 | 0.97 | 0.68 | 1.39 |
| Eastern | 1.17 | 0.80 | 1.71 | 1.06 | 0.73 | 1.55 |
| Western | 1.03 | 0.69 | 1.53 | 0.78 | 0.53 | 1.15 |
| Southern | 0.80 | 0.56 | 1.16 | 1.02 | 0.71 | 1.46 |
| **Wealth Index** |  |  |  |  |  |  |
| Poorest® |  |  |  |  |  |  |
| Poorer | 0.64*** | 0.45 | 0.91 | 0.79 | 0.56 | 1.11 |
| Middle | 0.73* | 0.51 | 1.05 | 0.94 | 0.65 | 1.34 |
| Richer | 0.89 | 0.61 | 1.30 | 1.22 | 0.84 | 1.77 |
| Richest | 0.99 | 0.66 | 1.49 | 1.33 | 0.89 | 1.99 |
| **Religion** |  |  |  |  |  |  |
| Hindu® |  |  |  |  |  |  |
| Non-Hindu | 1.05 | 0.82 | 1.36 | 1.09 | 0.85 | 1.40 |
| **Caste** |  |  |  |  |  |  |
| SC/ST |  |  |  |  |  |  |
| OBC | 1.05 | 0.82 | 1.34 | 1.00 | 0.79 | 1.28 |
| General | 1.12 | 0.83 | 1.51 | 1.17 | 0.87 | 1.57 |

**Source :** Authors' own calculation using 75th round of National Sample Survey data.
**Abbreviations:** LCI- Lower confidence interval; UCI- Upper confidence interval; SC- Schedule Caste; ST-Schedule Tribe; OBC-Other backward caste.
**Note:** No immunization is the reference category; 95% confidence interval in parentheses; significance level: \*\*\*significant at 1%, \*\*significant at 5%, \*significant at 10%; ® is
*reference category of independent variables; Non-Hindu includes Muslims, Christianity and other religion.*

Similarly, the children aged 21-28 months (0.50; p<0.01) & 29-36 months (0.35; p<0.01) are found to be significantly less likely to receive fully immunization than 12-20 months. Likewise, urban children are (1.26; p<0.1) found to be more likely to receive fully immunization compared to rural children. Children belonging to the North-Eastern regions (0.60; p<0.01) are significantly less likely to receive fully immunization compared to the Northern region.

## Discussion

Our paper studied the immunization coverage and pattern among children aged 12-36 months in India. India has achieved a significant improvement in vaccination coverage over the years due to its substantial policy and program initiatives. Despite this significant inequality persists in various socio-economic and marginalized communities. India is yet to achieve the fully immunization status with 2 percent of its child population is yet to be covered by any vaccination program as shown by our results. There is also a significant disparity in terms of vaccination coverage among the vulnerable group as shown by our results. Partial vaccination is still a challenge in India for policy makers, since a significant account of child is still receiving it, risking the health and wellbeing of children.

Multiple studies have been conducted in India so far to study the immunization coverage using various data sources reflecting the growth and challenges of immunization coverage in India. This study was a similar attempt to address the challenges of vaccination coverage in India and understand its differentials and determinants. The results where coherent with the results from earlier studies, but some of the results were contrary to some of the studies of this public health issue. Our results showed that 61 percent are fully immunized, while 37 are partially immunized as compared to 2 percent, which still lack any immunization coverage in India. This small but significant chunk of population mainly comes from rural areas, given the multiple underlying factors that affect the access and provision to immunization coverage among the rural children. These findings were consistent with earlier studies both in India and other countries [20–22]. The likely differences that have been attributed to this rural urban differential is mainly the access with greater concentration of health care facilities in urban areas as compared to rural areas. Similarly, supply-side factors like, availability, distance from the health center and associated to cost to access the healthcare facility [23].

One of the significant finding of this study was the contrary results that can be observed when analyzing the coverage at regional level. While our results showed that coverage is less likely in North East and Eastern regions of India, earlier findings have been contrary to this. A study [24] based on DHS data showed that vaccination coverage is lower in northern region of India, while as another showed that coverage is lower in the eastern region of India followed by Northeast [25]. The likely differences can be due to the different data set used in our study, apart from the timing of survey since this is the latest one in this context. Furthermore the difference can also be likely due to outreach programs and access to health care services in rural areas apart from the cost and waiting time associated with immunisation [26].

This study also examined the risk factors like age and sex of the children with respect to their vaccination coverage and found that age and sex significantly affects the vaccination status of children. Females children are more vaccinated as compared to the male children, although the differences are very less. Our findings were substantiated with some earlier findings from Nepal [27] and Nigeria [28]. But also contrary to various other studies [25,29–32]This may be attributed to the fact that the immunization programs not fully but partially have focused on gender perspective and stressed on the health of girl child in the recent years in India. Several girl specific program incentives have been launched, which are likely yielding the benefits as shown by our results.

The paper further studied the socio-economic determinants of immunization and the results confirm that socio-economic factors are key risk factors of immunization coverage coherent with the earlier studies in this context. Result indicate that lower concentration of immunization coverage is among the poor and marginalized groups in India. These findings complement with the findings reported by earlier studies in this context [24]. Similarly, the results found that full immunization is higher among upper wealth quintiles and urban areas coherent with the earlier studies [25,33]. Socio-economic characteristics are therefore key to understand the coverage of immunization, since these likely impact the poor coverage of immunization among children in India.

## Limitations

Like all other studies, the present study also has limitations. The study did not include all other variables due to unavailability of data. There may be other correlates that could capture the immunization with the data.

## Supporting information

Decription of variables

## Data Availability

The data analyzed are publicly available.

http://mospi.nic.in/unit-level-data-report-nss-75th-round-july-2017-june-2018-schedule-250social-consumption-health

## Declarations

### Ethics approval and consent to participate

Not applicable.

### Consent for publication

Not applicable.

### Competing interests

The authors declare that they have no competing interests.

### Funding

None.

## Acknowledgement

None

## Availability of data and materials

The data analyzed are publicly available. http://mospi.nic.in/unit-level-data-report-nss-75th-round-july-2017-june-2018-schedule-250social-consumption-health

## Notes

### Competing Interest Statement

The authors have declared no competing interest.

### Funding Statement

No funding

